# Pathogenic germline variants in a racially diverse real-world cohort of prostate cancer patients

**DOI:** 10.1101/2025.08.13.25333614

**Authors:** Taylor B Crawford, Maliha Tayeb, Emanuel Barrett, Tara Al-Saleem, Pranavi Kondam, Ryan Hausler, Heather Symecko, Caitlin Orr, Catherine Wolfe, Derek Mann, Kelsey Spielman, Gloria Wong, Naomi Haas, Vivek Narayan, Kyle Robinson, Samuel Takvorian, Yu-Ning Wong, Alexandra Sokolova, Robert B. Montgomery, Carolyn Menendez, Michael J. Kelley, Lisa B. Aiello, Isla P. Garraway, Susan M. Domchek, Kara N. Maxwell

**Author notes:** **Corresponding Author**: Kara N. Maxwell MD PhD, BRB II/III, 8th Floor. 421 Curie Blvd. Philadelphia, PA 19104.

## Abstract

**Importance:** Pathogenic germline variant (PGV) rates in cancer risk genes and their association with clinical pathological features inform genetic testing practices for prostate cancer (PCa) patients.

**Objective:** To determine the rate of PCa risk gene PGVs in a real-world, diverse cohort of PCa patients and identify clinical predictors of carrier status.

**Design, Setting, Participants, Main Outcomes and Measures:** Genetic testing results for 12 PCa risk genes, clinical, pathological, and family history of cancer variables were abstracted from clinical records for 1032 PCa patients who met National Comprehensive Cancer Network (NCCN) genetic testing criteria in oncology clinics and 3602 PCa patients who underwent testing in the VA National Precision Oncology Program (VA-NPOP). Individual gene PGV rates in PCa patients were compared to cancer-free males. Statistical testing was performed by unpaired t-tests and Fisher’s exact tests, corrected for multiple testing.

**Results:** Of 4634 PCa patients, 5.4% had PGVs in one of 12 PCa risk genes. PGVs in *BRCA2* (1.7%), *ATM* (1.3%), *CHEK2* (1.1%), Lynch genes (0.9%), and *BRCA1* (0.5%) were most common. The total PGV rate was significantly higher in 2825 self-identified White versus 1527 self-identified Black PCa patients (6.3% vs 3.7%, adjp=0.0024), although rates of *BRCA2* and *BRCA1* PGVs were similar (1.9% vs 1.3%, adjp=0.0885 and 0.6% vs 0.5%, adjp=0.8005, respectively). PGV rates were not significantly different in 311 Hispanic compared to 4188 non-Hispanic PCa patients (3.9% vs 5.5%, adjp=1.000). In self-identified White patients, PGV rates in *ATM*, *BRCA1*, *BRCA2, CHEK2* and Lynch genes were significantly higher compared to two cancer-free male cohorts. *BRCA2* and Lynch genes PGV rates were significantly higher in PCa patients compared to a cancer-free control cohort in SIRE-Black men. In a multivariable logistic regression, age at initial PCa diagnosis and self-identified race were significantly associated with any PGV.

**Conclusions and Relevance:** In a racially diverse, real-world cohort of individuals with PCa, lower PGV rates were identified compared to prior academic cohort studies. Outside of *ATM* and *CHEK2*, PGV rates were similar across the majority of clinical and pathological groupings. Our data support NCCN guideline indicated universal genetic testing for patients with aggressive forms of PCa.

**Key Points:** *Question:* What is the rate of pathogenic germline variants (PGVs) in a racially diverse, real-world cohort of prostate cancer patients who meet NCCN prostate cancer genetic testing criteria?

*Findings:* In this cohort study of 4634 prostate cancer patients who underwent genetic testing, we report a 5.4% PGV rate, lower than previously reported. No significant difference was seen in high-risk gene PGV rates by self-identified race or ethnicity. Age at initial PCa diagnosis, self-identified race, and prostate cancer in a first degree relative is significantly associated with having a PGV.

*Meaning:* Our data support current genetic testing guidelines of all patients with aggressive forms of prostate cancer.

## Introduction

Approximately 5% of all prostate cancers (PCa) are familial. Familial PCa is caused by a combination of non-genetic and genetic factors, including rare, dominantly inherited high/moderate risk variants and common low risk variants^1, 2^. For example, rare pathogenic germline variants (PGVs) in DNA repair genes such as *BRCA2* and the homeobox transcription factor *HOXB13* lead to increased risk of PCa^3,4^. Genetic testing of germline DNA is currently recommended by the National Comprehensive Cancer Network (NCCN)^5^ for males with a diagnosis of metastatic, regional (N1) or high risk/very high risk localized PCa. Genetic testing can identify patients, for example those with *BRCA1, BRCA2*, and Lynch Syndrome gene PGVs, who may benefit from FDA approved targeted therapy with PARP inhibitors and immune check point blockade, respectively^6^. PARP inhibitor indications are now expanding in PCa to include the neoadjuvant^7^ and metastatic hormone sensitive settings^8^, increasing the therapeutic relevance for early genetic testing. Germline genetic testing is also a critical part of clinical care because first-degree relatives of an affected carrier have a 50% chance of carrying the same genetic predisposition to developing cancer and either intervention or a higher level of surveillance can reduce cancer morbidity for family members^9^.

Reported PGV prevalence has varied between 9-17% in existing reports, depending on the population cohort and genes analyzed^10–13^. However, current studies have limited racially and ethnically diverse patient populations and are often reported from high-risk ascertained research or genetic testing company cohorts. Guidelines now recommend offering testing to all high risk/metastatic PCa patients, however, the prevalence in a real world, racially diverse, clinical cohort to inform genetic counseling practices and patient expectations remains to be elucidated. We report PGV rates and clinical-pathological associations of PGV status in a racially diverse cohort of PCa patients meeting NCCN genetic testing criteria.

## Methods

### Study Design

This is a retrospective cohort study of genetic testing results and clinical and pathological associations of rare PGVs in males with PCa. Inclusion criteria for the discovery cohort were males with a personal history of PCa who met National Comprehensive Cancer Network (NCCN) criteria for genetic testing due to PCa history (metastatic, regional (node positive), very high risk localized and high risk localized) and were seen in oncology clinics at one academic and three Veterans Affairs medical centers. Data were available on all patients offered genetic testing and the genetic testing results at Penn Medicine (n=864, genetic testing dates 1/1/2018-8/30/2023), Corporal Michael Crescenz VA Medical Center in Philadelphia (n=224), the Greater Los Angeles VA Medical Center (n=305), and the Puget Sound VA Medical Center in Seattle, WA (n=131) (all VA sites genetic testing dates 9/30/2021–7/5/2024). Inclusion criteria for the validation cohort were males with a diagnosis of PCa (identified by ICD billing codes) who underwent genetic testing for cancer risk genes based on NCCN criteria with the VA National Precision Oncology Program (VA-NPOP), excluding Veterans from the oncology clinic cohort (genetic testing dates 9/30/2021–7/5/2024). Patients were studied at Penn Medicine under a University of Pennsylvania Institutional Review Board approved, HIPAA-exempt protocol (#854011) and within the VA under a Central Institutional Review Board (CIRB) approved, HIPAA-exempt protocol entitled VA Multi-Omics Analysis Platform for Prostate Cancer and Sequencing (VA RESOLVE, CIRB#1742695).

### Data Collection

Patients underwent genetic testing using multiple clinical genetic testing panels. PGVs were studied in the following 12 genes tested in all patients: *ATM, BRCA1, BRCA2, CHEK2, EPCAM, HOXB13, NBN, MLH1, MSH2, MSH6, PMS2, TP53.* Genetic testing results including any identified PGVs were available for all cohorts; variants of uncertain significance (VUSs) were provided by all cohorts except Puget Sound (as VUS are not reported by their reference lab) and therefore those patients were excluded from VUS analyses. Clinical and pathological data and family history data were abstracted from the medical record at Penn Medicine and locally at the Philadelphia VAMC. Variables extracted included age at PCa diagnosis, self-reported race and ethnicity, vital status, clinical stage and risk category at diagnosis, biochemical recurrence, development of any metastatic PCa, and family history of breast, ovarian, pancreatic and prostate cancer. For all other VA patients, study participants were matched to variables in the VA-MAPP-Seq data repository which included age at PCa diagnosis, self-reported race and ethnicity, vital status from the Department of Defense, Gleason score, stage from the VA Cancer Registry data, and metastatic status from the Prostate Cancer Data Core. Clinical outcomes data for all patients was abstracted 7/30/2024.

### Comparison of PGV rates to cancer-free control males

PGV rates were determined per gene in each individual cohort, the combined discovery cohort, the validation cohort and the overall cohort. PGV rates in each gene from whole exome sequencing data from gnomAD and the Penn Medicine Biobank (PMBB) were obtained as previously described^14^. Briefly, all variants in the studied genes were annotated with ANNOVAR. Putative loss of function (pLOF) variants were identified using a combination of Clinvar pathogenicity calls, variant effect annotations, and splicing predictors. *CHEK2* p.I157T was excluded.

### Statistical analyses

Fisher’s exact test was used to compare PGV and VUS rates between SIRE-White and Black patients, self-identified Hispanic and non-Hispanic patients, and between the overall cohort versus cancer-free individuals in gnomAD and PMBB. An unpaired t-test comparing carriers and non-carriers was performed for age at initial diagnosis. Single variable association tests were completed by a Fishers exact test for all other clinical variables comparing all carriers vs non carriers and *BRCA2* carriers vs non-carriers. Multivariable logistic regression models to identify variables associated with any PGV and *BRCA2* PGVs were performed in the entire cohort using age at diagnosis, self-identified race, self-identified ethnicity, metastatic status, Gleason grade and vital status. Multivariable logistic regression models to identify variables associated with any PGV and *BRCA2* PGVs using all clinical variables were performed in the Penn Medicine and Philadelphia VA subset. Statistical tests were performed in GraphPad Prism or R. Hochberg corrected p-values were calculated for all Fishers exact tests performed.

## Results

Individuals with PCa who met NCCN genetic testing criteria (metastatic, regional, and high risk/very high risk localized PCa) were identified in oncology clinics at one academic (Penn Medicine) and three Veterans Affairs (VAMC) medical centers. Of 1526 oncology clinic patients approached, 1032 (68%) underwent genetic testing, 596 (69%) at the academic site, 171 (76%) at the Philadelphia VA, 155 (52%) at the Los Angeles VA, and 110 (82%) at the Puget Sound VA (**Fig.1**). The most common reason for not receiving genetic testing was active or passive declination.

**Figure 1.**
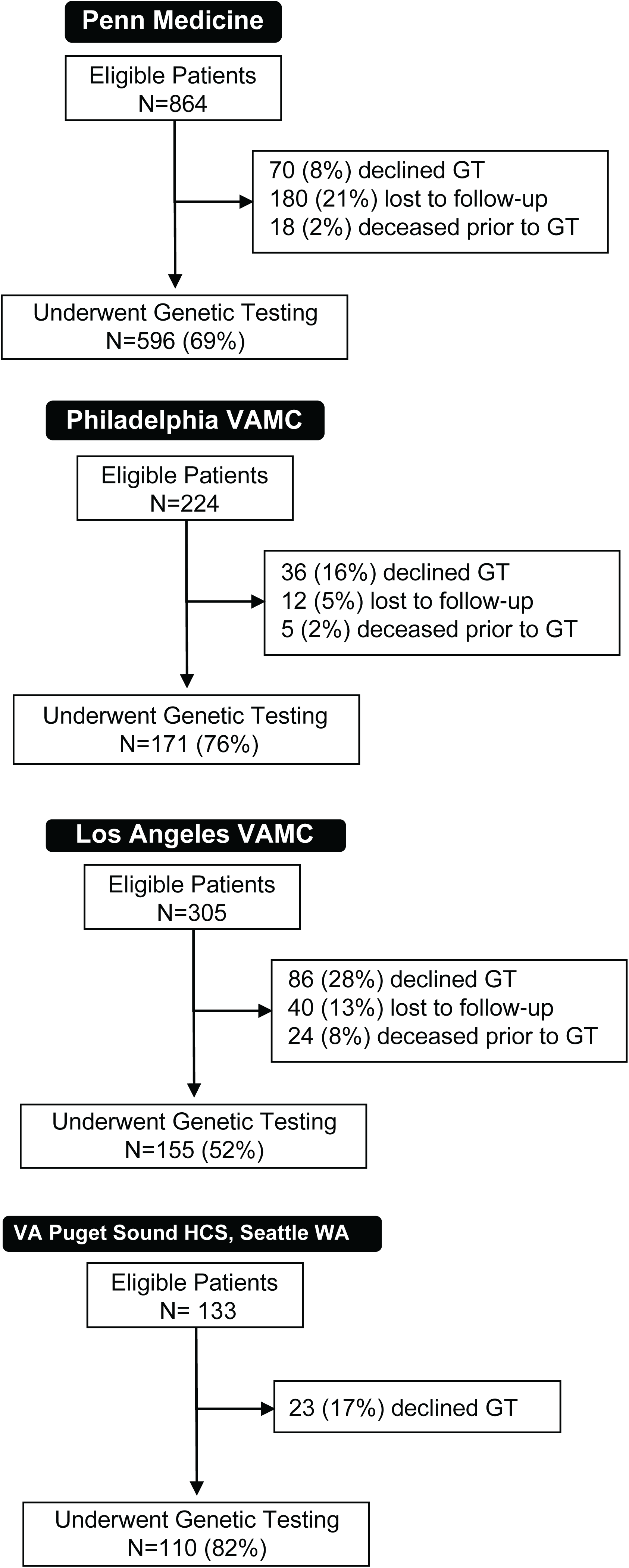
Consort diagrams of eligible prostate cancer patients for genetic testing at four Oncology Clinic sites. Males with prostate cancer who met NCCN guidelines for genetic testing (high risk and very high risk localized, regional N1M0, or metastatic M1) and presented to the respective medical oncology clinics at Penn Medicine, the Corporal Michael Crescenz Veterans Affairs Medical Center (Philadelphia VAMC), West Los Angeles Veterans Affairs Medical Center (Los Angele VAMC), and the VA Puget Sound Healthcare System (HCS) in Seattle, WA were tracked as eligible for genetic testing (“Eligible Patients”). Eligible patients who did not undergo genetic testing were categorized as having declining genetic testing, being lost to follow-up, or dying before testing could be conducted. GT, genetic testing.

The oncology clinic cohort (n=1032) included 654 (65%) self-identified White patients, 302 (30%) self-identified Black patients, and 32 (3%) self-identified Hispanic patients (**Table 1, Table S1**). The median(IQR) age of initial PCa diagnosis was 65(59-71). Most patients were metastatic at any point in their disease course (n=887, 86%). The majority of patients had Gleason Grade Group (GG) 4-5 PCa (n=541, 63%) and were initially diagnosed either localized or de novo metastatic (n=462, 47% and n=383, 39%, respectively) (**Table 1**). For validation, we evaluated genetic testing results of 3602 Veterans with a diagnosis of PCa and tested through the VA’s National Precision Oncology Program (VA-NPOP). This cohort included 2171 (62%) self-identified White patients, 1225 (35%) self-identified Black patients, and 279 (8%) self-identified Hispanic patients (**Table 1**). The median age of initial diagnosis was 67(60-73). Most patients were metastatic at any point in their disease course (n=2479, 69%) (**Table 1**). Most patients had GG4-5 PCa (n=1735, 55%) and were initially diagnosed with localized PCa (n=1159, 55%) (**Table 1**).

**Table 1:** Characteristics of the genetic testing cohort.

|  |  | **Oncology Clinical Cohort<sup>1</sup>** |  | **VA-NPOP** |  |
| --- | --- | --- | --- | --- | --- |
|  |  | **n** | **%** | **n** | **%** |
| **Age at Initial Diagnosis** |  | **(n=1032)** |  | **(n=3602)** |  |
|  | Age (IQR) | 65(59-71) |  | 67 (60-73) |  |
| **Self-reported Race, n (%)** |  | **(n=1002)** |  | **(n=3474)** |  |
|  | White | 654 | 65% | 2171 | 62% |
|  | Black | 302 | 30% | 1225 | 35% |
|  | Asian | 11 | 1% | 14 | 0.4% |
|  | Other | 35 | 3% | 64 | 2% |
| **Self-reported Ethnicity, n (%)** |  | **(n=1013)** |  | **(n=3486)** |  |
|  | Hispanic | 32 | 3% | 279 | 8% |
| **Ashkenazi Jewish Status, n (%)** |  | **(n=643)** |  | NA |  |
|  | Yes | 51 | 8% |  |  |
| **Vital Status, n (%)** |  | **(n=1032)** |  | **(n=3602)** |  |
|  | Alive | 824 | 80% | 3253 | 90% |
| **Metastatic at any point, n (%)** |  | **(n=1032)** |  | **(n=3602)** |  |
|  | Yes | 887 | 86% | 2479 | 69% |
| **Clinical Stage at Initial Diagnosis, n (%)** |  | **(n=983)** |  | **(n=2100)** |  |
|  | Localized | 462 | 47% | 1159 | 55% |
|  | Regional | 138 | 14% | 429 | 20% |
|  | De novo metastatic | 383 | 39% | 512 | 24% |
| **Gleason Grade, n (%)** |  | **(n=866)** |  | **(n=3139)** |  |
|  | GG1 | 62 | 7% | 305<sup>4</sup> | 10% |
|  | GG2 | 101 | 12% | 570 | 18% |
|  | GG2/3 | 59 | 7% | N/A | N/A |
|  | GG3 | 103 | 12% | 493 | 3% |
|  | GG4 | 178 | 21% | 654 | 21% |
|  | GG5 | 363 | 42% | 1081 | 34% |
| **T Stage, n (%)** |  | **(n=360)** |  |  |  |
|  | T1 | 81 | 23% |  |  |
|  | T2 | 83 | 23% | NA |  |
|  | T3 | 183 | 51% |  |  |
|  | T4 | 13 | 4% |  |  |
| **N Stage, n (%)** |  | **(n=585)** |  | **(n=2100)** |  |
|  | N1 | 177 | 30% | 162 | 8% |
| **Risk Stage at Initial Diagnosis<sup>2</sup>, n (%)** |  | **(n=418)** |  |  |  |
|  | Very Low/Low | 48 | 11% | NA |  |
|  | Intermediate | 131 | 31% |  |  |
|  | High/Very High | 239 | 57% |  |  |
| **Family History of Cancer, n (%)<sup>3</sup>** |  | **(n=767)** |  |  |  |
|  | Prostate Cancer | 297 | 39% |  |  |
|  | Breast Cancer | 194 | 25% | NA |  |
|  | Ovarian Cancer | 35 | 5% |  |  |
|  | Pancreatic Cancer | 40 | 5% |  |  |

<sup>1</sup>The oncology clinical cohort includes Penn Medicine (n=596), Philadelphia VAMC (n=171), Greater Los Angeles VAMC (n=155), and VA Puget Sound HCS (n=110).
<sup>2</sup>Risk stage at initial diagnosis for patients diagnosed with localized PCa, VA Puget Sound HCS did not report localized risk stage at initial diagnosis.
<sup>3</sup>Greater Los Angeles VAMC and VA Puget Sound HCS did not report family history of cancer.
NA, not available; PCa, prostate cancer

Of 1032 PCa patients meeting NCCN criteria for genetic testing in Oncology Clinics, 7.1% (n=73) were found to have a PGV in at least one of 12 genes (**Fig. 2a, Table S2-3**). PGVs were most frequently found in *BRCA2* (n=20, 1.9%), *ATM* (n=17, 1.6%), *CHEK2* (n=12, 1.2%), and *BRCA1* (n=11, 1.1%) (**Fig.2a, Table S2-3**). Overall, 13.2% of patients were found to have at least one or more VUS. VUS were most common in *ATM* (n=31, 3.4%), *BRCA2* (n=18, 2.0%), *PMS2* (n=18, 2.0%), and *MSH6* (n=14, 1.5%) (**Fig. S1a, Table S2-S3**). Of 3602 Veterans with PCa tested by VA-NPOP, 4.9% (n=178) were found to have a PGV in at least one of 12 genes (**Fig. 2a, Table S4**). PGV rates were most common in *BRCA2* (n=59, 1.6%), *ATM* (n=44, 1.2%), and *CHEK2* (n=37, 1.0%). Overall, 18.3% of patients were found to have at least one or more VUS. VUS were most common in *ATM* (n=180, 5.0%), *MSH6* (n=83, 2.3%), *BRCA2* (n=78, 2.2%), and *PMS2* (n=63, 1.7%) (**Fig.S1a, Table S4**). Given that VA-NPOP patients could have been tested for an indication other than PCa, a sensitivity analysis of 2473 patients who had testing specifically with a prostate cancer panel were evaluated. Like the overall cohort, 4.2% (n=105) of patients had a PGV, and 19.4% had a VUS (**Table S5**).

**Figure 2.**
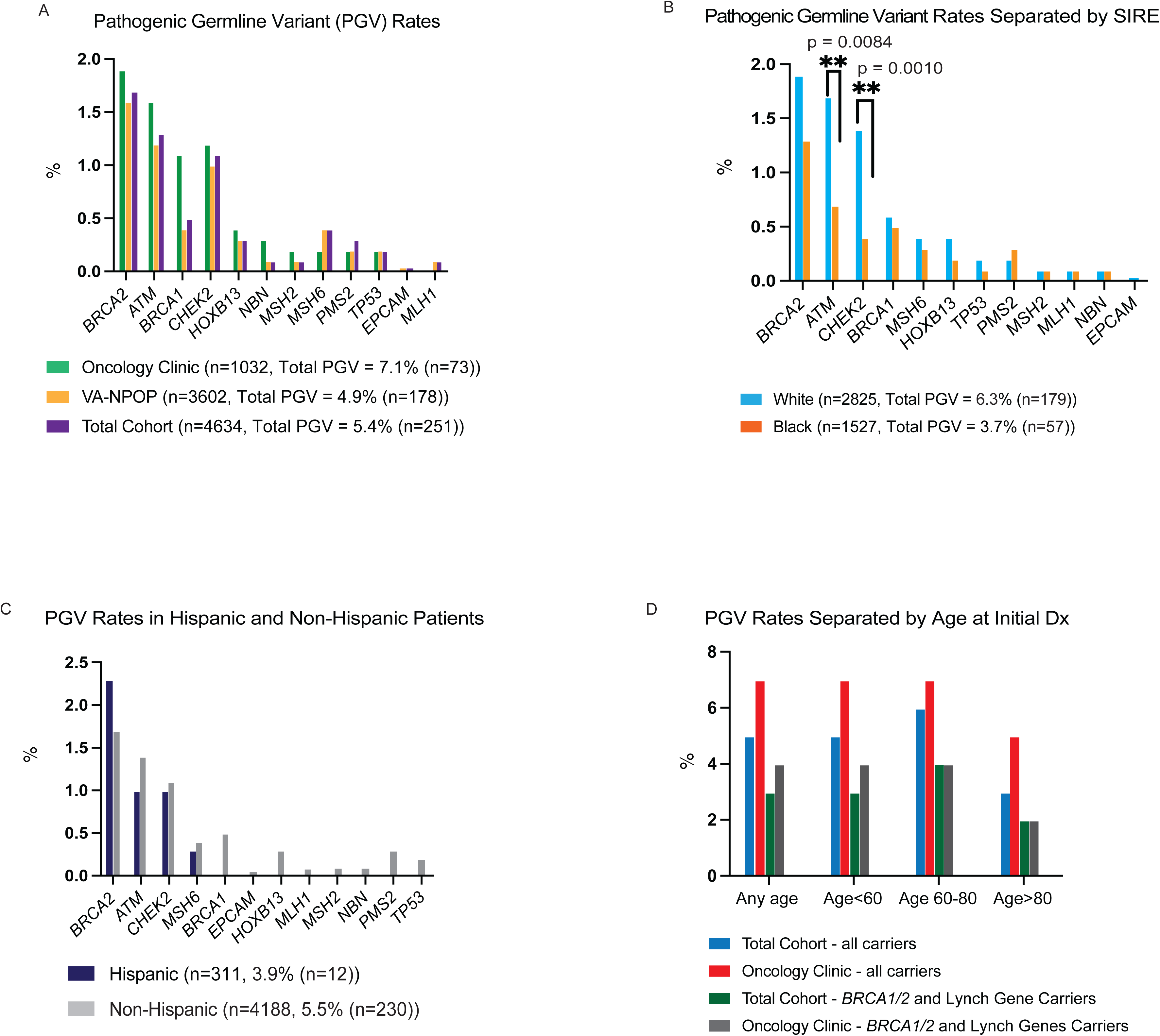
Rates of pathogenic germline variants and variants of uncertain significance in males with prostate cancer eligible for genetic testing. (A) Pathogenic germline variants (PGV) rates for individual genes in the Oncology Clinic cohort (n=1,032), VA National Precision Oncology Program cohort (NPOP, n=3,602), and total cohort combined (n=4,634). (B) PGV rates in total combined cohort for individual genes stratified by self-identified race/ethnicity (SIRE) White (n=2,825) and Black (n=1,527). (C) PGV rates in Hispanic (n=311) and Non-Hispanic (n=4188) patients with PCa. (D) PGV rates separated by age groups at initial prostate cancer diagnosis.

Combining the Oncology Clinic and VA-NPOP cohorts (n=4634), 5.4% (n=251) of patients had a PGV (**Table S6**). We stratified PGV and VUS rates by self-identified race and ethnicity in the combined cohort. Self-identified White patients (n=2825) had a significantly higher rate of PGVs compared to self-identified Black patients (n=1527) (6.3% vs 3.7%, adjp=0.0024). However, the difference in PGV rates was driven by differences in *ATM* and *CHEK2* PGVs (1.7% vs 0.7%, adjp=0.0504 and 1.4% vs 0.4%, adjp=0.0070, respectively, in self-identified White and Black patients) (**Fig.2b, Table S6**). It is notable that self-identified White and Black patients had similar PGV rates in *BRCA2* (1.9% vs 1.3%, adjp=0.0885), *BRCA1* (0.6% vs 0.5%, adjp=0.8005), and the Lynch genes (0.9% vs 0.9%, adjp=1.000) (**Fig.2b, Table S6**). PGV rates were also similar in self-identified Hispanic compared to non-Hispanic patients overall (3.9%, vs 5.5%, adjp=1.000) and in each gene (**Fig.2c, Table S6**). The VUS rates in self-identified White patients were nominally lower than self-identified Black patients (15.8% vs 19.8%, adjp=0.0568) (**Fig. S1b, Table S6**). VUS rates between Hispanic and Non-Hispanic patients were similar (18.1% vs 17.2%, adjp=1.000). (**Fig.S1c, Table S6**). Finally, we stratified PGV rates by age at diagnosis. Including all genes, PGV rates were 5.8%, 5.5% and 3.3% if diagnosed age<60, age 60-80, and age>80, respectively, although this was not statistically significant (**Fig. 2d, Table S7**). Similarly, PGV rates were similar for *BRCA1, BRCA2, MLH1, MSH2, MSH6, PMS2* across age at diagnosis categories.

The combined cohort was next used to determine whether PGVs were significantly enriched among PCa patients compared to two cancer-free male cohorts from gnomAD and the Penn Medicine Biobank (PMBB)^14^. In genes with at least three carriers per group, we found that PGV rates were significantly higher in the advanced PCa patients compared to both gnomAD and PMBB for *BRCA2*, *BRCA1*, *ATM, CHEK2,* and in the Lynch syndrome genes (*MLH1, MSH2, MSH6,* and *PMS2*) in self-identified White patients (**Table 2**). Low numbers of self-identified Black patients precluded statistical testing; however, PGV rates were nominally higher in *BRCA2* and the Lynch syndrome genes (**Table 2**).

**Table 2:** Comparison of PGV rates in the clinical PrCa cohort versus cancer-free males.

| Gene | Genetic Testing Cohort |  | gnomAD Cancer-Free Males |  |  | PMBB Cancer-Free Males |  |  |
| --- | --- | --- | --- | --- | --- | --- | --- | --- |
|  | White, n=2825 |  | White, Avg n=11750 |  |  | White, n=2615 |  |  |
|  | n | % | n | % | p-value<sup>1</sup> | n | % | p-value |
| ***BRCA2*** | 54 | 1.91% | 38 | 0.30% | <0.0001 | 14 | 0.50% | <0.0001 |
| ***ATM*** | 47 | 1.67% | 44 | 0.40% | <0.0001 | 16 | 0.60% | 0.0003 |
| ***BRCA1*** | 17 | 0.60% | 30 | 0.30% | 0.008 | 5 | 0.20% | 0.02 |
| ***CHEK2*** | 40 | 1.41% | 83 | 0.70% | 0.0005 | 7 | 0.30% | <0.0001 |
| **Lynch<sup>2</sup>** | 25 | 0.88% | 43 | 0.40% | 0.001 | 4 | 0.20% | 0.0001 |
|  | Black, n=1527 |  | Black, Avg n=1450 |  |  | Black, n=515 |  |  |
|  | n | % | n | % | p-value | n | % | p-value |
| ***BRCA2*** | 20 | 1.31% | 4 | 0.30% | 0.002 | 2 | 0.40% | na |
| **Lynch** | 13 | 0.85% | 3 | 0.20% | 0.02 | 0 | 0.00% | na |
| ***ATM*** | 11 | 0.72% | 6 | 0.40% | 0.33 | 0 | 0.00% | na |
| ***BRCA1*** | 7 | 0.46% | 3 | 0.20% | 0.34 | 2 | 0.40% | na |
| ***CHEK2*** | 6 | 0.39% | 3 | 0.20% | 0.51 | 1 | 0.20% | na |

<sup>1</sup>Fisher's exact testing performed if all comparator cells had n<sub>≥</sub>3 patients
<sup>2</sup>PGVs in *MLH1*, *MSH2*, *MSH6*, *PMS2* combined

We next tested the association of clinical and pathological features with PGV carrier status in the overall cohort of 251 carriers versus 4383 non-carriers. Age at initial diagnosis was similar in carriers compared to non-carriers [65(60-71) vs 66(60-73), respectively, adj.p=0.5235] (**Fig.3a; Fig.S2a; Table S8**). There was not a statistically significant difference in clinical stage at initial diagnosis, Gleason grade, and metastatic diagnosis at any point (**Fig. 3b, Fig.S2b-c; Table S8**) in PGV carriers versus non-carriers. For those patients initially diagnosed with localized PCa, clinical risk stage, T stage, and N stage were not statistically different between carriers and non-carriers (**Fig.S3a-c; Table S8**). Comparing 79 PCa patients with PGVs in *BRCA2* compared to 4555 non-*BRCA2* carriers, median age of diagnosis was similar [65(59-71) vs 66(60-72), adj.p=1.000] (**Fig.3a; Fig.S2a; Table S9)**. *BRCA2* carriers had higher Gleason grade group (GG4-5) compared to non-carriers (adj.p=0.0036, **Fig.3b, Table S9**). However, *BRCA2* carriers did not have enrichment for any other clinical or pathological variable compared to non-*BRCA2* carriers (**Fig S2a-c, S3a-c; Table S9**).

**Figure 3.**
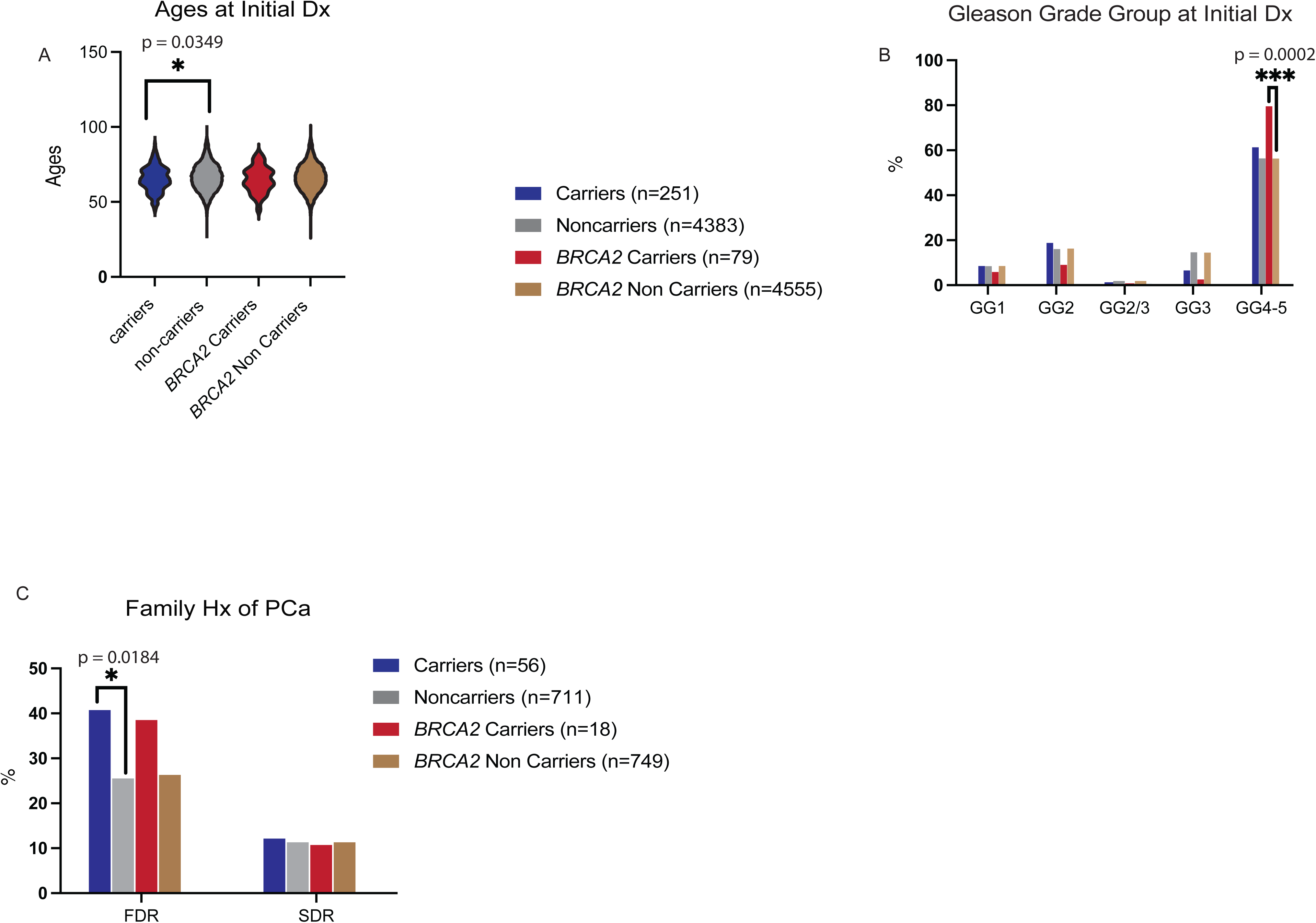
Analysis of clinical and pathological associations with PGV status in prostate cancer patients from the total cohort. **(A)** Age groups at initial prostate cancer diagnosis, **(B)** Fraction of patients with indicated Gleason grade group (GG) at initial diagnosis, **(C)** Fraction of patients with family history of prostate cancer, all variables shown in all PGV carriers, all PGV non-carriers, *BRCA2* PGV carriers, and *BRCA2* PGV non-carriers. Age distributions in carriers versus non-carriers and *BRCA2* carriers versus non-carriers each compared by a student’s T Test. Rates of each Gleason grade group or family history category in carriers versus non-carriers and *BRCA2* carriers versus non-carriers were compared by a Fisher’s exact test.

For the subset of patients with family history data (n=56 PGV carriers vs n=711 non-carriers), carriers were numerically more likely than non-carriers to have a first-degree relative with PCa (41% vs 26%, unadjp=0.0184, adj.p=0.2944) (**Fig.3c, Table S8**). There was no significant difference in family history individually of breast cancer, ovarian cancer, and pancreatic cancer between carriers and non-carriers (**Fig.S4a-c, Table S8**); however, carriers were more likely to have a first or second degree relative for any of these cancers compared to non-carriers (93% vs 72%, unadjp=0.0004, adjp=0.0072). *BRCA2* carriers were numerically more likely than non-carriers to have a first-degree relative with ovarian cancer (17% vs 3%, unadjp=0.0271, adjp=0.4336) and a first or second degree relative with any cancer (100% vs 73%, unadj.p=0.0056, adj.p=0.0952) (**Fig.S4b, Table S9**).

Finally, we used multivariable logistic regression to identify variables associated with any PGV and *BRCA2* PGVs. All clinical variables were available for the Penn Medicine and Philadelphia VA subset. We found that only age at initial diagnosis and self-identified race were associated with any PGV (**Table S10**). We therefore repeated the analysis in the entire cohort using variables available in >70% of patients, namely, age at diagnosis, self-identified race, self-identified ethnicity, metastatic status, Gleason grade and vital status. We found that age at initial PCa diagnosis and self-identified race were significantly associated with any PGV, and age at initial PCa diagnosis and Gleason grade were significantly associated with a *BRCA2* PGV (**Table S10**).

## Discussion

We report the rate of PGVs in DNA repair PCa risk genes to be 5.4% in a racially diverse cohort of 4634 PCa patients, including 1527 self-identified Black men (33%), who met NCCN personal history criteria for genetic testing and consented to genetic testing practices. The PGV rate was largely contributed to by *BRCA2* (1.7%), *ATM* (1.3%), *CHEK2* (1.1%) and *BRCA1* (0.5%) and PGVs in these genes were found at significantly higher rates in the high risk localized and advanced PCa cohort compared to cancer-free males. Additionally, age at initial diagnosis and self-identified race were significantly associated with finding any PGV in a multivariable logistic regression model. *BRCA2* PGV status was significantly associated with age at initial diagnosis and high Gleason grade at diagnosis and notably not associated with self-identified race.

Reported PGV prevalence has widely varied in existing reports and the prevalence of PGVs in a real-world diverse cohort of high risk/advanced prostate cancer patients remains uncertain. Prior published studies have shown 9-17% PGV rates^10–13^; however, cohorts consisted of either no^10^ or under 6% self-identified Black patients^12–13^. These studies reported higher PGV rates than our study likely due to inclusion of genes with less robust evidence of association with PCa, more ‘selected’ patient cohorts (such as academic studies and genetic testing companies), and minimal inclusion of self-identified Black and Hispanic patients. Our study reports on individuals seen in clinical settings and is the largest cohort of self-identified Black (n=1527) and Hispanic patients (n=311). For example, a recent genetic testing study called GENTleMEN reported 9% of patients carried a PGV.^11^ However, their study included the low risk *CHEK2* I157T variant; if these carriers are removed, the GENTleMEN study found a PGV rate of 7.2%, similar to our study. The most identified genes in our study are similar to prior studies, namely *BRCA2*, *ATM, CHEK2,* and *BRCA1*. It is important to note that our data strongly suggest that *BRCA1* and *BRCA2* PGV rates do not differ by self-identified race and ethnicity in PCa patients.

*BRCA1/2* mutations are associated with aggressive prostate cancer phenotypes, overall poorer survival, and early onset of disease^15–23^; however, data is less robust for other DNA repair genes. Despite the large cohort size, PGV rarity, sample size, and limited follow-up time precluded study of associations of clinical and pathological features by individual gene outside of *BRCA2*. We did find evidence in this study that *BRCA2* was associated with higher Gleason grade, consistent with prior studies. Overall, however, only age at PCa diagnosis and family history of any of breast, ovary, pancreas and/or prostate cancer was associated with carriage of a PGV.

The limitations for our study include lack of complete data for all patients and the maximum follow-up time from genetic testing was six years. The comparison of PGV rates was not made to an identically ascertained cohort of cancer-free males; however, comparisons were made to both gnomAD and an academic biobank with similar results.

In conclusion, we report that approximately 5% of males with PCa who meet NCCN genetic testing criteria have a PGV in a PCa risk gene. Although we found a significant difference between carriers and non-carriers for age at initial diagnosis and self-identified race, most clinical and pathological variables included in this study do not predict carrier status in a cohort of advanced PCa patients. Further, self-identified race and ethnicity were not associated with PGVs in high-risk genes that have important clinical implications for treatment. Therefore, our work supports the current NCCN guideline based recommendation for testing all males with aggressive PCa to inform clinical treatment decisions and familial testing.

## Data Availability Statement

It is not possible for the authors to directly share the individual-level data that were obtained from the patients included in this study due to constraints stipulated in the informed consent. Anyone wishing to gain access to this data should inquire directly to the authors.The data generated from our analyses are included in the manuscript main text, tables, and figures and online Supplementary Materials (available online).

