## Supplemental Figures for "Pathogenic germline variants in a racially diverse real-world cohort of prostate cancer patients"

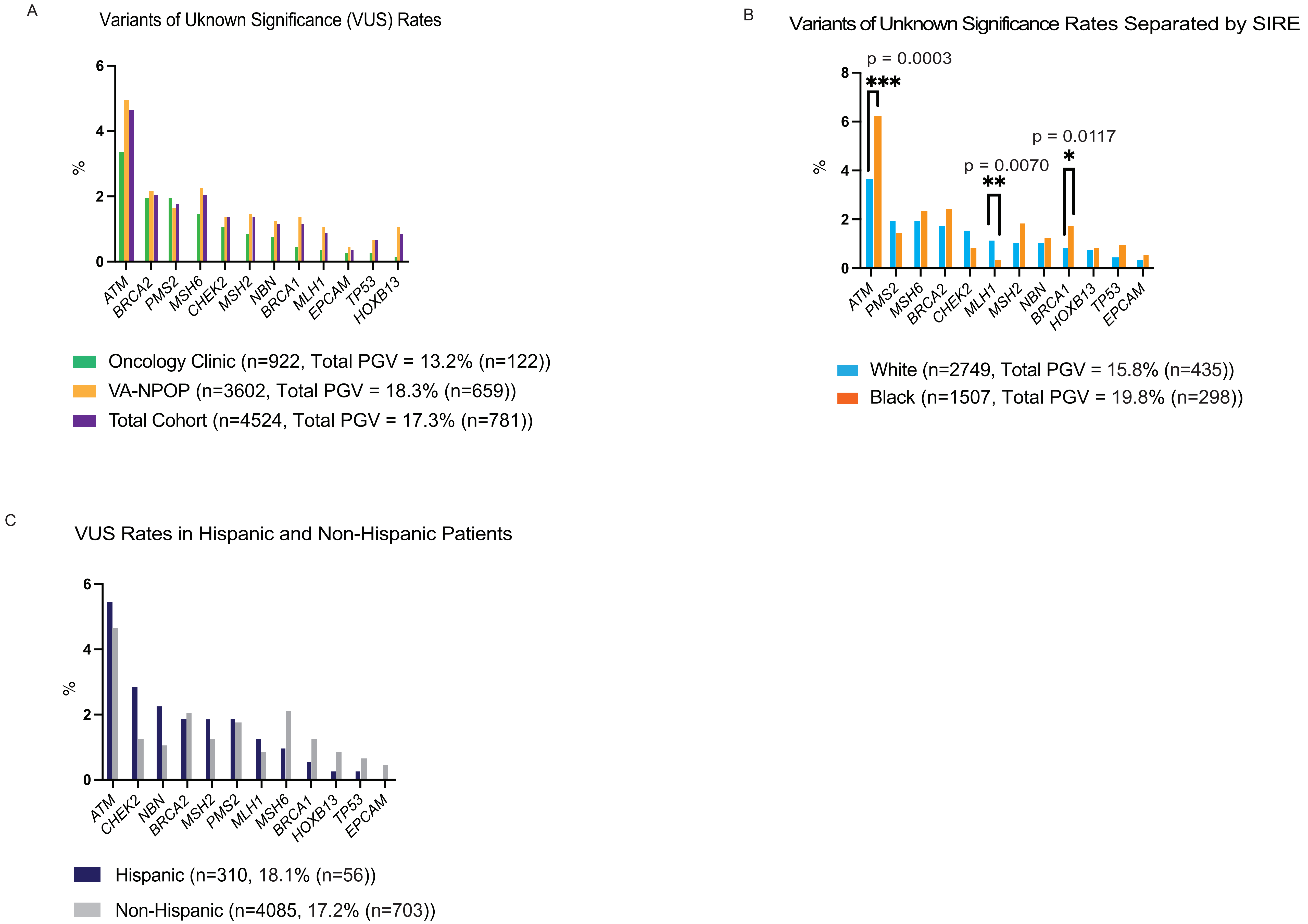


**Supplemental Figure 1:** Rates of variants of uncertain significance (VUS) in males with prostate cancer. (A) VUS rates in oncology clinic, VA-NPOP, and total cohorts for individual genes. (B) VUS rates stratified by White and Black self-identified ethnicity. (C) VUS rates in Hispanic and Non-Hispanic patients.

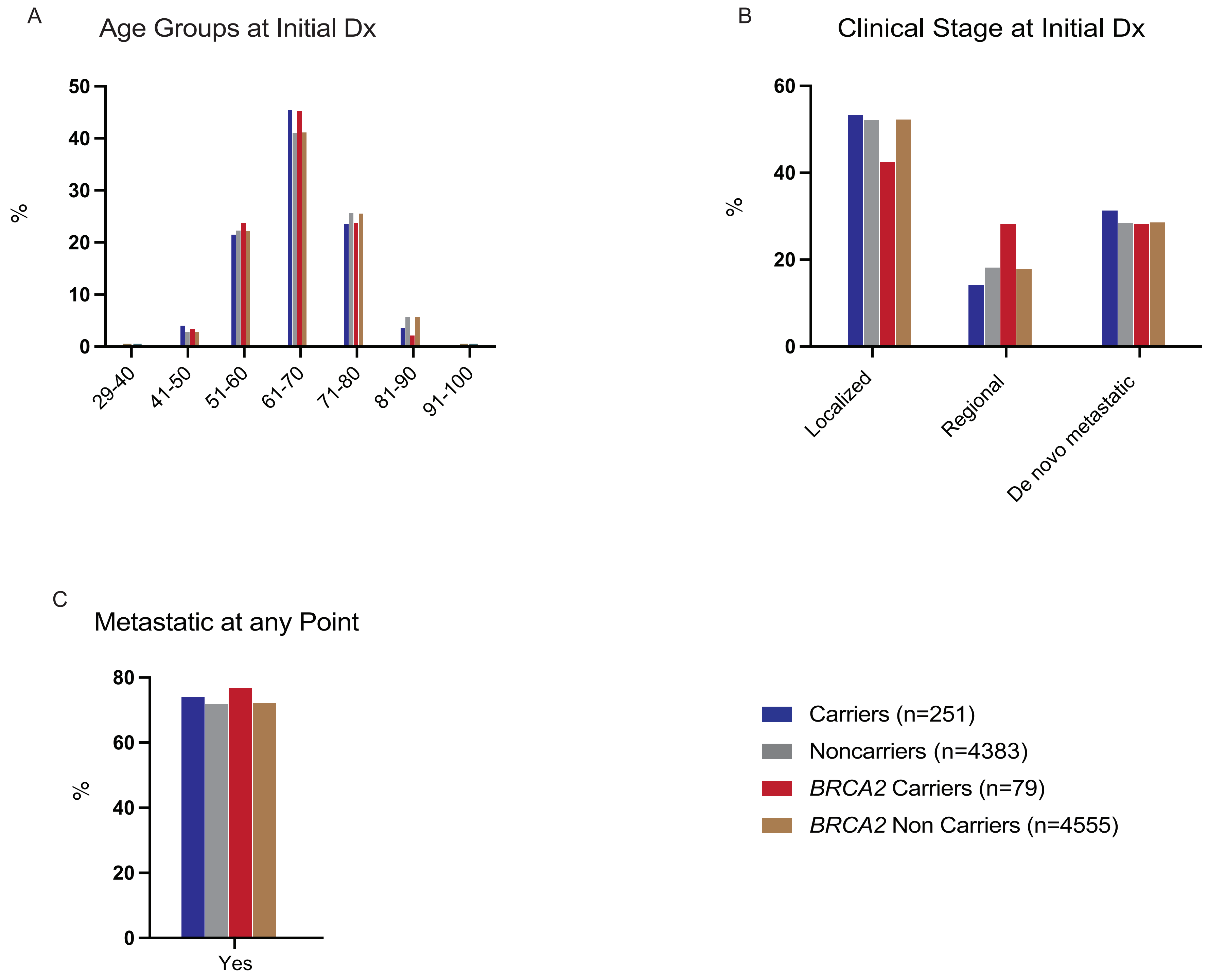


**Supplemental Figure 2.** Analysis of clinical and pathological associations with PGV status in prostate cancer patients from the total cohort. (A) Age groups at initial prostate cancer diagnosis, (B) fraction of patients with indicated broad clinical stage at initial diagnosis, and (C) fraction of patients who become metastatic at any point in their disease course. All variables shown in all PGV carriers, all PGV non-carriers, BRCA2 PGV carriers, and BRCA2 PGV non-carriers

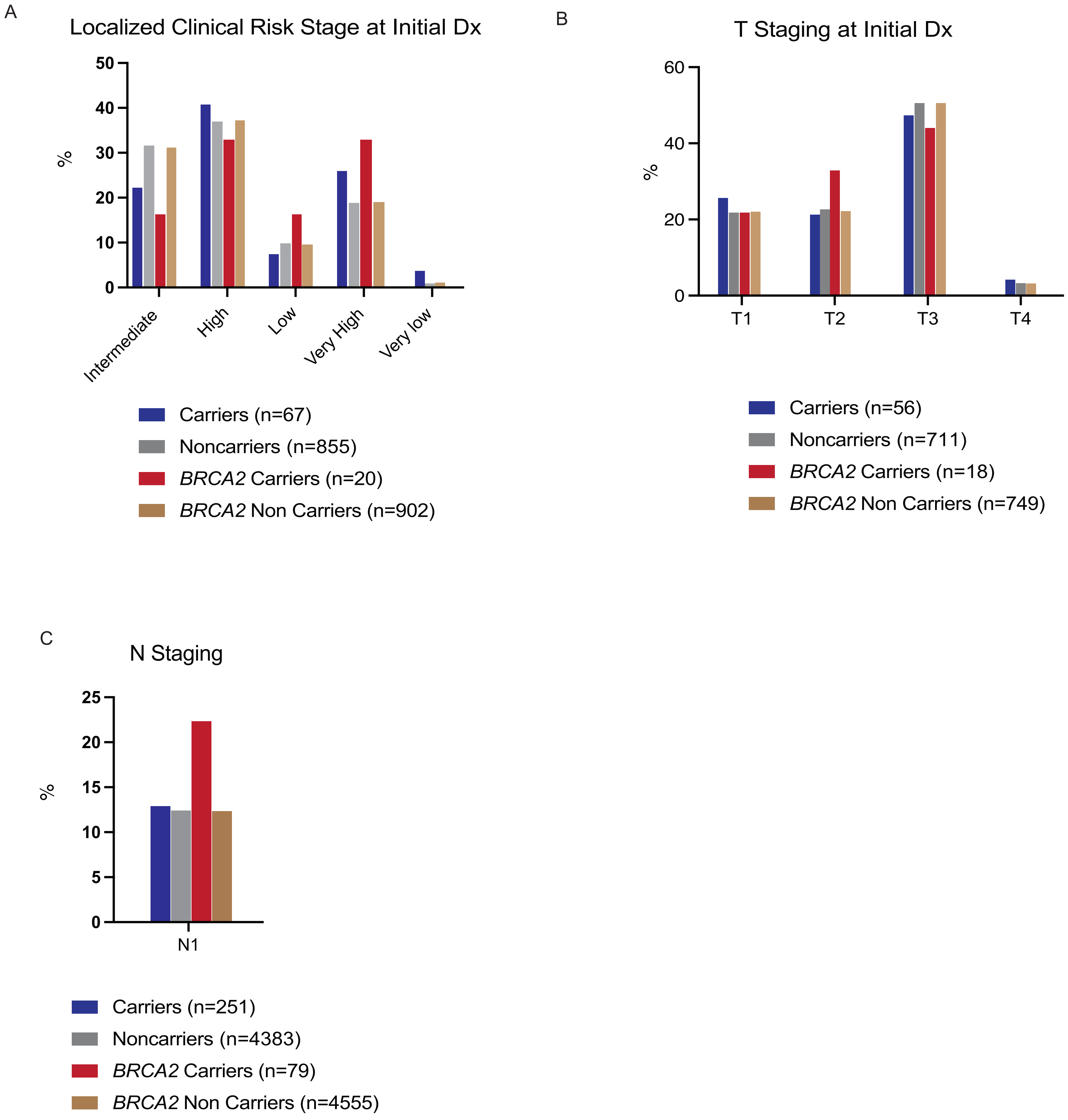


**Supplemental Figure 3:** Analysis of clinical and pathological associations with PGV status in prostate cancer patients from the total cohort. (A) Fraction of patients with indicated localized clinical risk stage at diagnosis. (B) Fraction of patients with indicated T stage at initial diagnosis. (C) Fraction of patients with N1 stage at initial diagnosis. Variables shown in all PGV carriers, all PGV non-carriers, BRCA2 PGV carriers, and BRCA2 PGV non-carriers

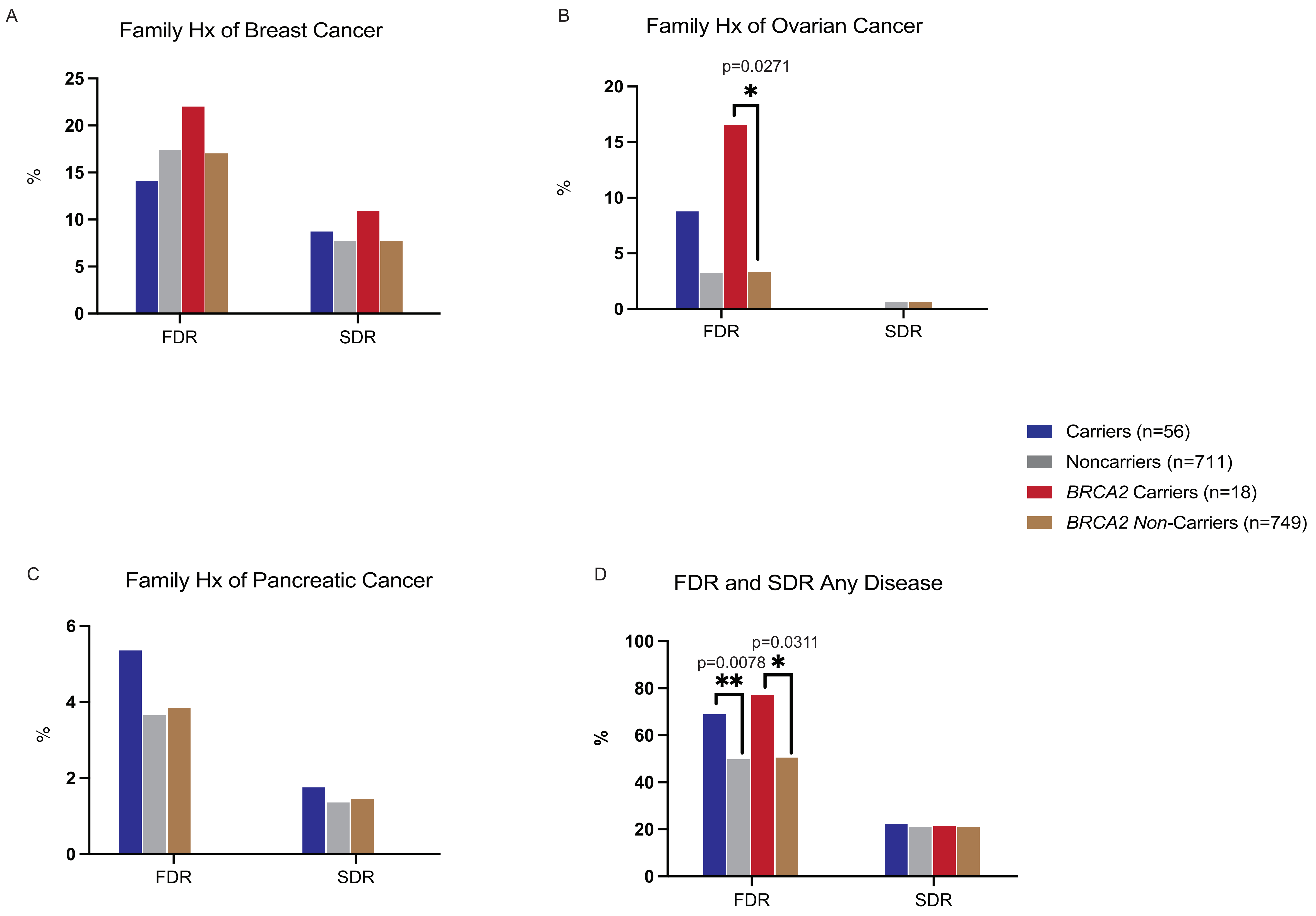


**Supplemental Figure 4:** Analysis of family history associations with PGV status in prostate cancer patients from Penn Medicine and Philadelphia VAMC. Fraction of patients with at least one (A) FDR and SDR with breast cancer, (B) FDR and SDR with ovarian cancer, (C) FDR and SDR with any cancer, (D) Any family history of any cancer in all PGV carriers, all PGV non-carriers, BRCA2 PGV carriers, and BRCA2 PGV non-carriers with available family history data Penn Medicine and the Philadelphia VAMC. FDR, first-degree relative; SDR, second-degree relative.
